# Epidemiology and Antimicrobial Resistance of Klebsiella pneumoniae at a Tertiary Oncology Centre in Astana, Kazakhstan, 2022–2026: A Prospective Longitudinal Surveillance Study

**DOI:** 10.64898/2026.10.23.26366297

**Authors:** Gulfairus A. Yessenbayeva, Irina Y. Mukatova, Yelena V. Zholdybayeva, Yevgeniy A. Merenkov, Yevgeniy S. Avakyan, Gulbanu S. Berdiyarova, Sanzhar B. Shalekenov, Makhira S. Khudaibergenova, Zhandos A. Burkitbayev

## Abstract

**Background:** *Klebsiella pneumoniae* is a World Health Organization (WHO) critical priority pathogen, and carbapenem-resistant *K. pneumoniae* (CRKP) causes 50,000–100,000 attributable deaths annually worldwide. Systematic antimicrobial resistance (AMR) data from Central Asia are virtually absent from the global literature.

**Methods:** Longitudinal surveillance study of routine microbiological data from the National Research Oncology Centre (NROC), Astana, Kazakhstan (January 2022–March 2026). Four data sources were integrated: monthly microbial landscape reports (October 2022–September 2024; n=596 *K. pneumoniae* isolates), a full-year patient-level clinical register (2025; n=91 isolates), monthly landscape reports for first quarter 2026 (Q1 2026 n=50 isolates), and biannual antibiograms (2022, 2026). Isolates were classified as CRKP (4MRGN), 3MRGN, ESBL-producing, or susceptible.

**Results:** A total of 737 *K. pneumoniae* isolates were identified across the full study period. CRKP (4MRGN) was the dominant phenotype throughout, peaking at 76.4% in January–September 2024 and remaining at 62.0% in Q1 2026. The aggregate MDR rate (CRKP + 3MRGN + ESBL) was 76–87% across all periods, consistently 2–3 times above the global pooled mean of 28.7%. The highest resistance burden was observed in the bone marrow transplantation department (86–98% MDR) and intensive care units (67–100% MDR). Between 2022 and 2026, a profound shift in reserve antibiotic susceptibility was documented: amikacin resistance rose from 8.5% (2022) to 38.1% (Q1 2026, whole institution) and 60% (Q1 2026, intensive care sector), nearly eliminating the sole previously active antibiotic class. In contrast, polymyxins (colistin R 0%, polymyxin B R 2%) and tigecycline (R 52%) emerged as the only agents with retained activity in 2026. Carbapenem resistance showed a modest partial improvement (meropenem R 72%→64%) but remained critically high. Simultaneously, *Escherichia coli* (ESBL+/MDR) emerged as the dominant pathogen in haematological and surgical departments by 2025–2026, reflecting a broader Enterobacterales resistance crisis beyond *K. pneumoniae* alone.

**Conclusions:** This study provides the first systematic published AMR surveillance evidence from Kazakhstan over a four-year period. CRKP dominates the institutional microbiological landscape, and the progressive loss of aminoglycoside activity now leaves polymyxins and tigecycline as the last remaining active agents — a development with profound implications for patient safety. These findings represent a critical call to action for antimicrobial stewardship implementation, carbapenemase genotyping, and integration into national and global AMR surveillance systems in Central Asia.

## 1. Introduction

*Klebsiella pneumoniae* is a Gram-negative, encapsulated opportunistic pathogen and a leading cause of healthcare-associated infections worldwide, including bloodstream infections (BSIs), pneumonia, urinary tract infections, and surgical site infections^[1]^ and has been described as a major worldwide source and shuttle for antibiotic resistance genes, facilitating horizontal transfer to other Enterobacterales^[2]^

Its clinical burden is disproportionately concentrated in immunocompromised patients — particularly those undergoing haematological malignancy treatment or haematopoietic stem cell transplantation (HSCT) — in whom bacteraemia carries mortality rates of 30–50%.^[3,4]^

The WHO has classified carbapenem-resistant *K. pneumoniae* as a “critical priority pathogen”.^[5]^ The Global Burden of Disease (GBD) 2019 AMR analysis estimated CRKP caused 50,000–100,000 attributable deaths globally in a single year,^[4]^ and the 2024 update confirmed CRKP among the fastest-growing AMR mortality threats, with attributable burden increasing substantially from 1990 to 2021.^[6]^ In Europe alone, AMR caused an estimated 33,000 attributable deaths in 2015.^[7]^ CRKP infections carry pooled all-cause mortality of 42.1% versus 21.2% for susceptible strains,^[8]^ with an odds ratio for death in BSI of approximately 3.87.^[9]^

Despite this global urgency, AMR surveillance data from Central Asia are almost entirely absent from published literature. A Kazakhstani ICU study noted its data were, to the authors knowledge, the only published report from a Central Asian country, with epidemiological data “underrepresented and poorly reported”.^[10]^ A subsequent analysis from the same institution documented rapidly increasing carbapenem resistance rates in 2015, with resistance in ICU-related HAIs already exceeding European benchmarks threefold. ^[11]^ A 2025 WHO report from Uzbekistan similarly described the need to strengthen basic AMR diagnostics and national surveillance capacity.^[12]^ GBD analyses document increasing *K. pneumoniae* burden among elderly populations in Eastern Europe and Central Asia.^[13]^

The National Research Oncology Centre in Astana is Kazakhstan’s principal tertiary cancer centre, caring for patients undergoing intensive chemotherapy, haematopoietic stem cell transplantation (HSCT), and complex surgical procedures — a population at extreme risk for MDR infections. No systematic published study has characterised *K. pneumoniae* AMR epidemiology at NROC or any comparable Kazakhstani institution.

This study describes the prevalence, temporal trends, departmental distribution, and antibiotic susceptibility of *K. pneumoniae* at NROC over a four-year period (January 2022–March 2026), with particular attention to the evolution of reserve antibiotic activity — a dimension that has transformed substantially during the study period and carries direct and urgent implications for clinical practice.

## 2. Materials and Methods

### 2.1 Study Design and Setting

Longitudinal surveillance study of routinely collected microbiological data from NROC, Astana, Kazakhstan — a 350-bed tertiary cancer centre. Microbiological analyses are performed in the institutional Clinical Diagnostic Laboratory using automated culture systems (BacT/ALERT, bioMérieux) and standardised susceptibility testing in accordance with national standards and European Committee on Antimicrobial Susceptibility Testing (EUCAST) breakpoints.

### 2.2 Data Sources

#### Source 1 — Monthly microbial landscape reports (October 2022–September 2024)

Standardised monthly reports presenting organism counts by department, specimen type, and resistance phenotype. Twenty monthly datasets covering 596 *K. pneumoniae* isolates.

#### Source 2 — Patient-level clinical microbiology register, full-year 2025

Patient-identified register of all clinically significant isolates, January 1–December 31, 2025 (n=91 *K. pneumoniae* isolates). This source captures only clinically significant isolates, not routine surveillance cultures, and is not directly comparable in denominator to landscape reports.

#### Source 3 — Monthly landscape reports and antibiograms, Q1 2026 (January–March 2026)

Full institutional landscape data (12 files; two parts per month for landscape and antibiogram respectively) covering all departments, January–March 2026 (n=50 *K. pneumoniae* isolates for landscape; n=50 for antibiogram). This source provides the most current data and includes the first whole-institution antibiogram since 2022.

#### Source 4 — Annual antibiogram, 2022

Cumulative susceptibility data for 2022, institution-wide (meropenem n=159) and haemato-oncological departments (meropenem n=118). Together with Source 3, this enables direct 2022 vs. 2026 antibiogram comparison.

#### Source 5 — Institutional AMR monitoring report (2019–2023)

Annual comparative antibiotic susceptibility data for 2019–2023 providing longitudinal context.

### 2.3 Isolate Classification

**4MRGN (CRKP):** non-susceptible to carbapenems, 3rd-generation cephalosporins, fluoroquinolones, and aminoglycosides.

**3MRGN:** non-susceptible to three of four antibiotic classes, with retained carbapenem susceptibility.

**ESBL+:** ESBL-producing, resistant to 3rd-generation cephalosporins, carbapenem-susceptible.

**Susceptible:** not meeting the above criteria.

Aggregate MDR = 4MRGN + 3MRGN + ESBL+. *K. oxytoca* isolates excluded throughout.

### 2.4 Data Extraction and Analysis

Data were stored in institutional Microsoft Excel files (.xlsx/.xlsm format). Data extraction and aggregation were performed using Python 3 (openpyxl library, version 3.1). Standardised algorithms mapped resistance phenotype columns from monthly tabular reports, identified departmental subgroups from sheet headers, and summed isolate counts by period, department, and specimen type. Antibiogram resistance rates were calculated as R/(R+S) × 100%, where R denotes the number of resistant isolates and S the number of susceptible isolates. Due to heterogeneity in data sources across study periods, year-on-year comparisons are presented descriptively without formal statistical testing.

### 2.5 Ethical Considerations

This study was approved by the Local Ethics Committee of the National Centre for Biotechnology LLP, Astana, Kazakhstan (approval date: October 8, 2025). Data were anonymised prior to analysis. Individual patient consent was waived under national regulatory provisions for retrospective/prospective analysis of anonymised administrative and laboratory records.

## 3. Results

### 3.1 Overview: 2022–2026

Across the full study period (October 2022–March 2026), 737 *K. pneumoniae* isolates were identified across all data sources. CRKP (4MRGN) was the dominant phenotype throughout, with aggregate MDR rates of 76–87% across all periods — consistently 2–3 times above the global pooled mean of 28.7%.^[12]^ The complete period overview is presented in Table 1.

**Table 1.**
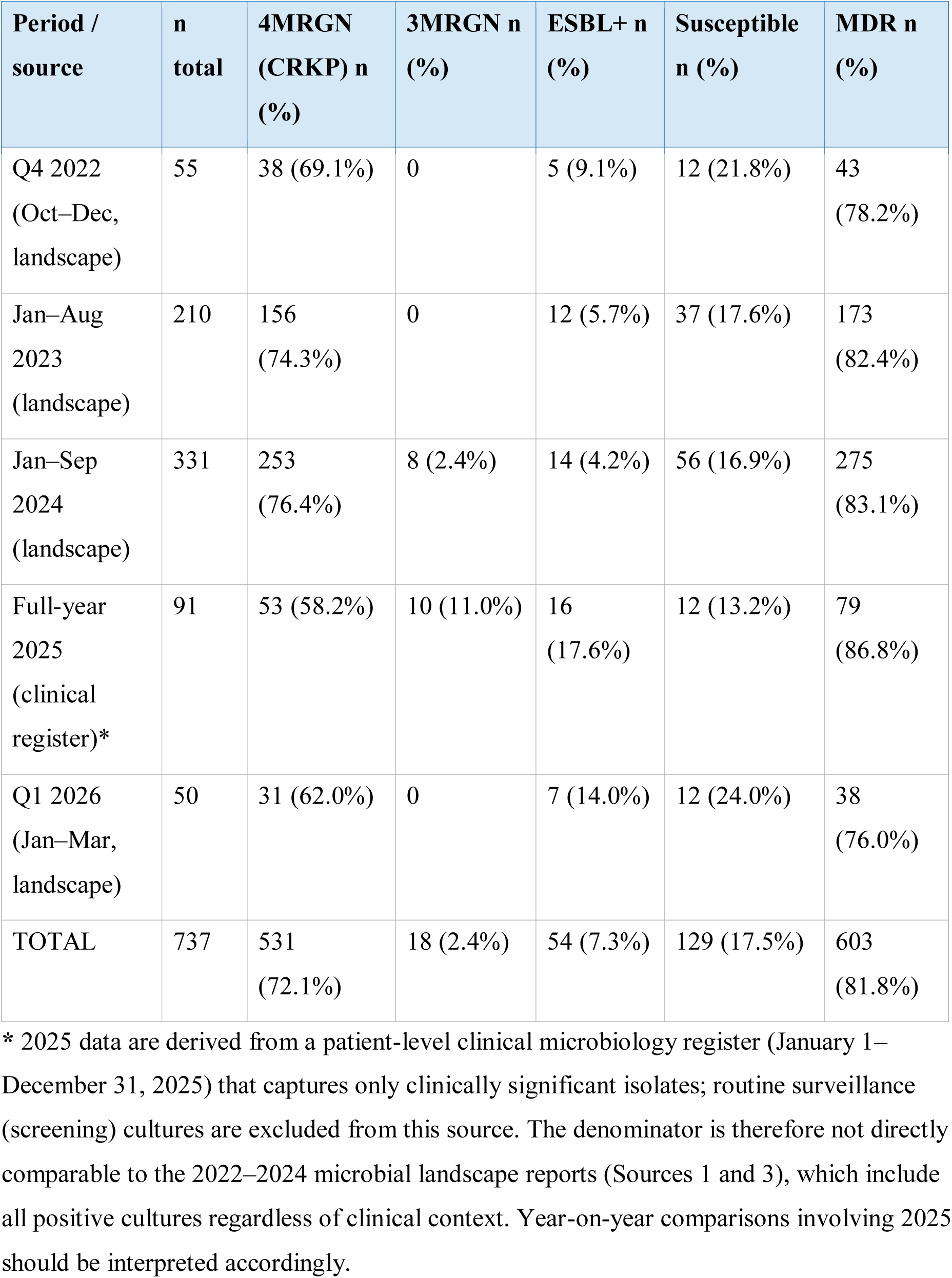
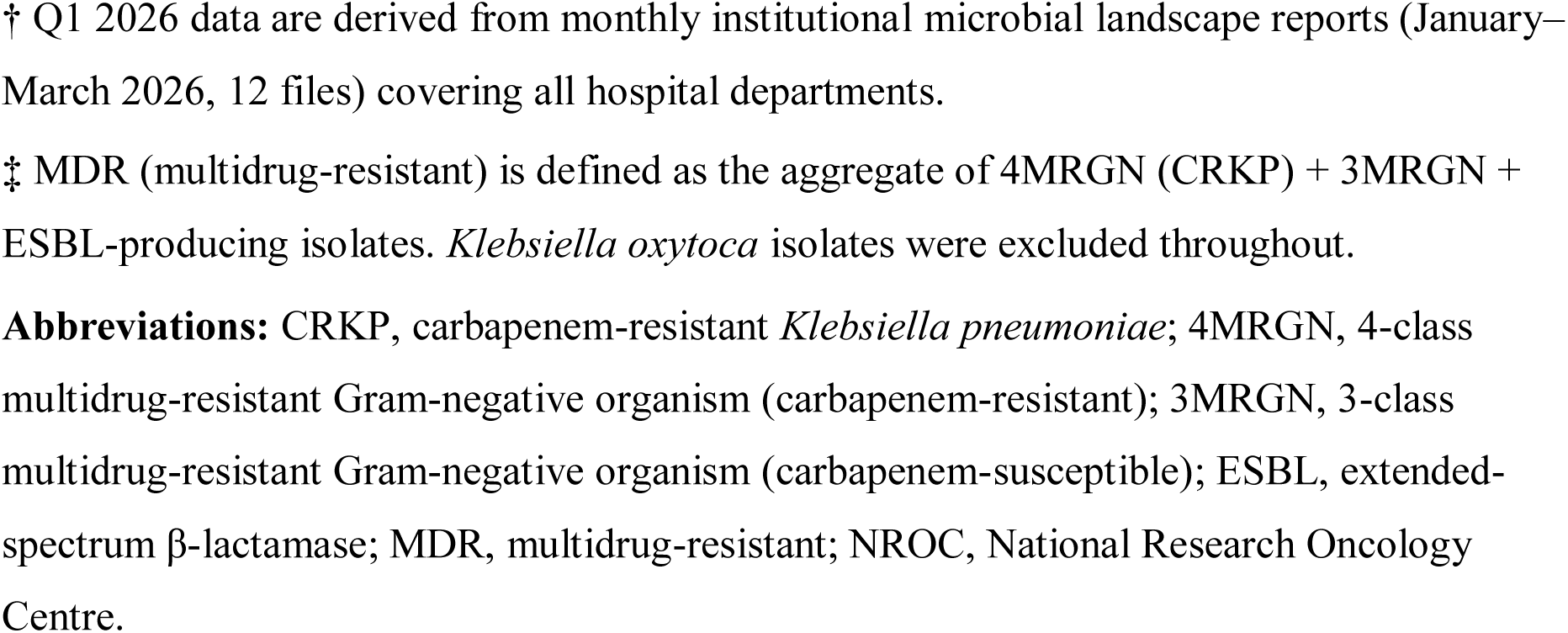
Overview of Klebsiella pneumoniae isolates by period and resistance phenotype, National Research Oncology Centre (NROC), Astana, Kazakhstan, 2022–2026.

| Period / source | n total | 4MRGN (CRKP) n (%) | 3MRGN n (%) | ESBL+ n (%) | Susceptible n (%) | MDR n (%) |
| --- | --- | --- | --- | --- | --- | --- |
| Q4 2022 (Oct–Dec, landscape) | 55 | 38 (69.1%) | 0 | 5 (9.1%) | 12 (21.8%) | 43 (78.2%) |
| Jan–Aug 2023 (landscape) | 210 | 156 (74.3%) | 0 | 12 (5.7%) | 37 (17.6%) | 173 (82.4%) |
| Jan–Sep 2024 (landscape) | 331 | 253 (76.4%) | 8 (2.4%) | 14 (4.2%) | 56 (16.9%) | 275 (83.1%) |
| Full-year 2025 (clinical register)* | 91 | 53 (58.2%) | 10 (11.0%) | 16 (17.6%) | 12 (13.2%) | 79 (86.8%) |
| Q1 2026 (Jan–Mar, landscape) | 50 | 31 (62.0%) | 0 | 7 (14.0%) | 12 (24.0%) | 38 (76.0%) |
| TOTAL | 737 | 531 (72.1%) | 18 (2.4%) | 54 (7.3%) | 129 (17.5%) | 603 (81.8%) |

\* 2025 data are derived from a patient-level clinical microbiology register (January 1–December 31, 2025) that captures only clinically significant isolates; routine surveillance (screening) cultures are excluded from this source. The denominator is therefore not directly comparable to the 2022–2024 microbial landscape reports (Sources 1 and 3), which include all positive cultures regardless of clinical context. Year-on-year comparisons involving 2025 should be interpreted accordingly.
† Q1 2026 data are derived from monthly institutional microbial landscape reports (January–March 2026, 12 files) covering all hospital departments.
‡ MDR (multidrug-resistant) is defined as the aggregate of 4MRGN (CRKP) + 3MRGN + ESBL-producing isolates. *Klebsiella oxytoca* isolates were excluded throughout.
**Abbreviations:** CRKP, carbapenem-resistant *Klebsiella pneumoniae*; 4MRGN, 4-class multidrug-resistant Gram-negative organism (carbapenem-resistant); 3MRGN, 3-class multidrug-resistant Gram-negative organism (carbapenem-susceptible); ESBL, extended-spectrum β-lactamase; MDR, multidrug-resistant; NROC, National Research Oncology Centre.

### 3.2 Temporal Trends: 2022–2024

CRKP was the dominant *K. pneumoniae* phenotype in all 20 available monthly landscape reports (October 2022–September 2024). Monthly proportions in 2024 ranged from 53.3% (June) to 97.8% (January and March). No sustained reduction in CRKP proportion was observed at any point in this period. Monthly data for 2024 are presented in Table 2.

**Table 2.**
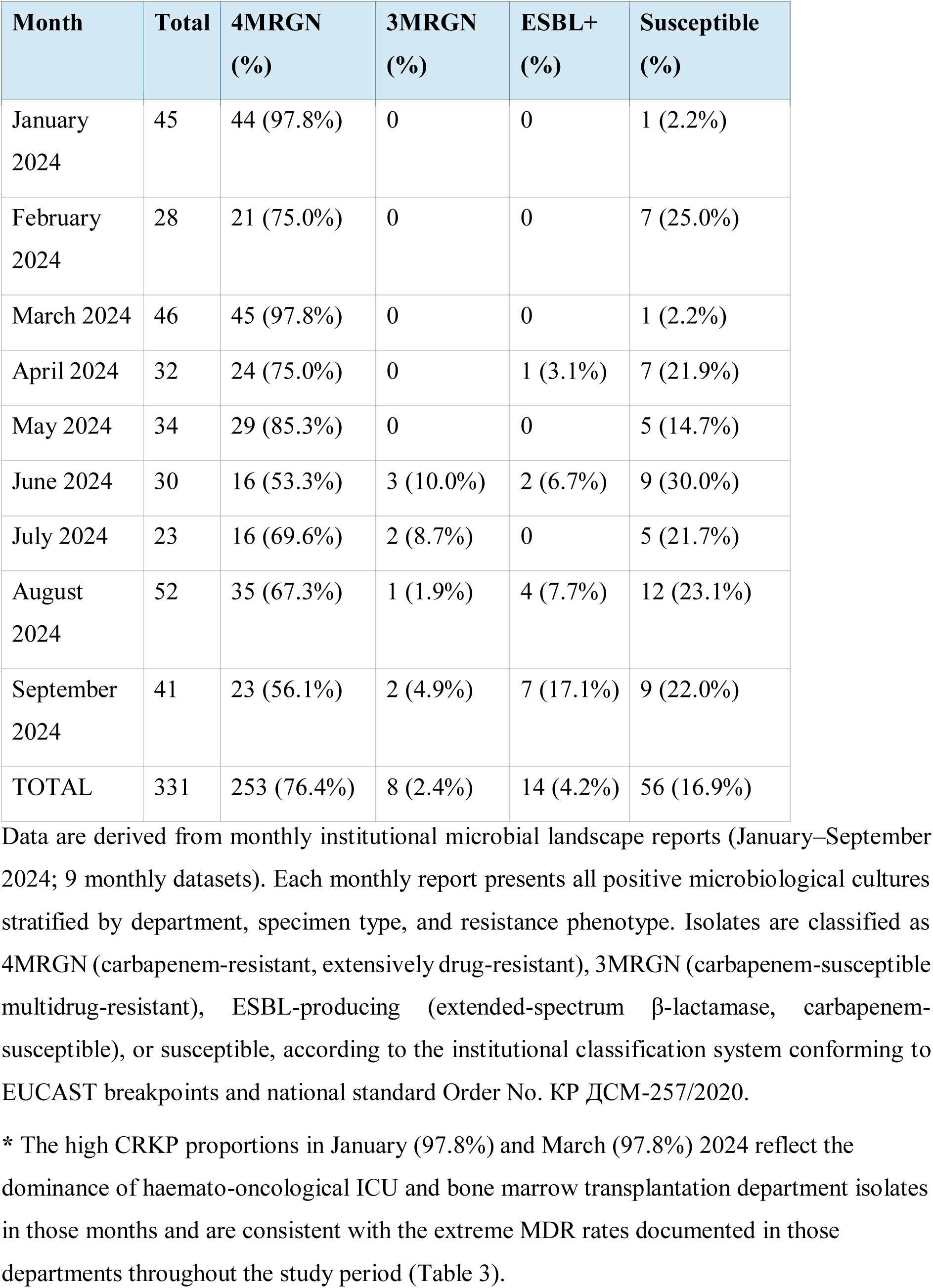
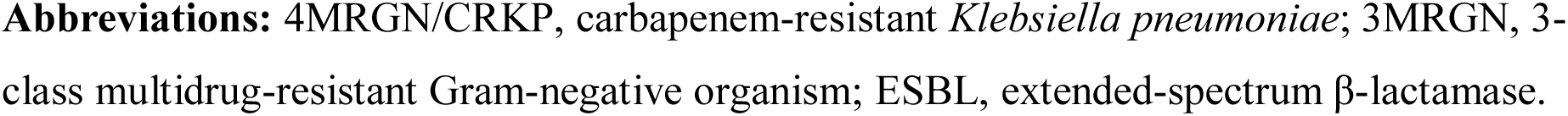
Monthly Klebsiella pneumoniae isolate counts by resistance phenotype, NROC Astana, January–September 2024.

| Month | Total | 4MRGN (%) | 3MRGN (%) | ESBL+ (%) | Susceptible (%) |
| --- | --- | --- | --- | --- | --- |
| January 2024 | 45 | 44 (97.8%) | 0 | 0 | 1 (2.2%) |
| February 2024 | 28 | 21 (75.0%) | 0 | 0 | 7 (25.0%) |
| March 2024 | 46 | 45 (97.8%) | 0 | 0 | 1 (2.2%) |
| April 2024 | 32 | 24 (75.0%) | 0 | 1 (3.1%) | 7 (21.9%) |
| May 2024 | 34 | 29 (85.3%) | 0 | 0 | 5 (14.7%) |
| June 2024 | 30 | 16 (53.3%) | 3 (10.0%) | 2 (6.7%) | 9 (30.0%) |
| July 2024 | 23 | 16 (69.6%) | 2 (8.7%) | 0 | 5 (21.7%) |
| August 2024 | 52 | 35 (67.3%) | 1 (1.9%) | 4 (7.7%) | 12 (23.1%) |
| September 2024 | 41 | 23 (56.1%) | 2 (4.9%) | 7 (17.1%) | 9 (22.0%) |
| TOTAL | 331 | 253 (76.4%) | 8 (2.4%) | 14 (4.2%) | 56 (16.9%) |

Data are derived from monthly institutional microbial landscape reports (January–September 2024; 9 monthly datasets). Each monthly report presents all positive microbiological cultures stratified by department, specimen type, and resistance phenotype. Isolates are classified as 4MRGN (carbapenem-resistant, extensively drug-resistant), 3MRGN (carbapenem-susceptible multidrug-resistant), ESBL-producing (extended-spectrum β-lactamase, carbapenem-susceptible), or susceptible, according to the institutional classification system conforming to EUCAST breakpoints and national standard Order No. KP ДСМ-257/2020.
\* The high CRKP proportions in January (97.8%) and March (97.8%) 2024 reflect the dominance of haemato-oncological ICU and bone marrow transplantation department isolates in those months and are consistent with the extreme MDR rates documented in those departments throughout the study period (Table 3).
**Abbreviations:** 4MRGN/CRKP, carbapenem-resistant *Klebsiella pneumoniae*; 3MRGN, 3-class multidrug-resistant Gram-negative organism; ESBL, extended-spectrum β-lactamase.

### 3.3 Epidemiological Shift: 2025–2026

The 2025 register and Q1 2026 landscape data (Table 3) documented a significant redistribution within the MDR spectrum. The proportion of 4MRGN among *K. pneumoniae* isolates declined from 76.4% (2024) to 62.0% (Q1 2026), but this was accompanied by a parallel rise in ESBL-producing strains (4.2%→14.0%) and, in 2025, in 3MRGN isolates (0→11.0%). The aggregate MDR rate remained 76–87% across both years, confirming that the apparent reduction in CRKP does not represent clinical improvement but a redistribution of resistance burden within the MDR spectrum.

**Table 3.** Klebsiella pneumoniae isolates by clinical department: 2025 and Q1 2026, NROC Astana.

| Department | 2025<br>Total | 2025 MDR<br>(%) | Q1 2026<br>Total | Q1 2026<br>MDR (%) |
| --- | --- | --- | --- | --- |
| Bone marrow transplant | 18 | 17 (94%) | 22 | 19 (86%) |
| Intensive care sector | —* | — | 10 | 8 (80%) |
| Anaesth./resuscitation<br>ICU | 24 | 21 (88%) | 9 | 6 (67%) |
| Haemato-oncology | 11 | 4 (36%) | 3 | 3 (100%) |
| Hepatopancreaticobiliary | 9 | 8 (89%) | — | — |
| Other departments | 29 | 27 (93%) | 6 | 2 (33%) |

— Indicates department not separately identifiable in the respective data source owing to institutional reclassification of unit names between 2024 and 2026, or absence of isolates in that period.
\* Intensive Care Sector data from the 2025 clinical register could not be separately identified due to departmental reclassification; the Q1 2026 figure is from the full institutional landscape report.
**Abbreviations:** MDR, multidrug-resistant.

Concurrently, institutional analytical reports documented a broader pathogen transition: *Escherichia coli* (ESBL+, MDR) displaced *K. pneumoniae* 4MRGN as the dominant organism in the bone marrow transplantation centre (; *E. coli* 52% in 2025 vs. *K. pneumoniae* 4MRGN ∼55% in 2024) and surgical ICU. In Q1 2026, bone marrow transplantation centre remained the highest-burden department for *K. pneumoniae* (MDR 86%, n=22 isolates), followed by the intensive care sector (MDR 80%, n=10) and the anaesthesiology/resuscitation centre (MDR 67%, n=9).

### 3.4 Antibiotic Susceptibility: 2022 vs. 2026

Direct comparison of the 2022 and Q1 2026 institutional antibiograms reveals a complex picture of partial carbapenem recovery alongside progressive loss of reserve aminoglycoside activity (Table 4).

**Table 4.**
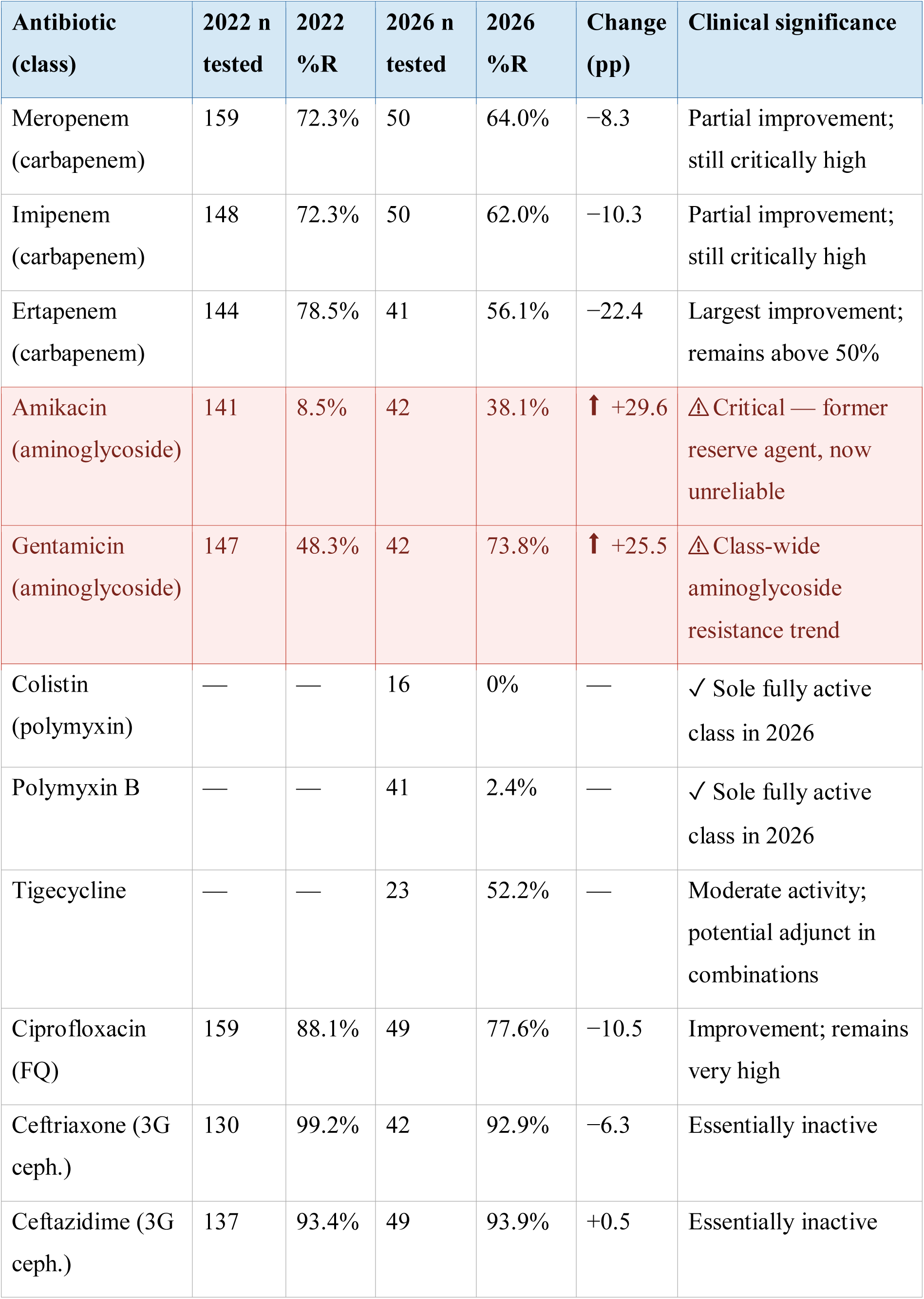
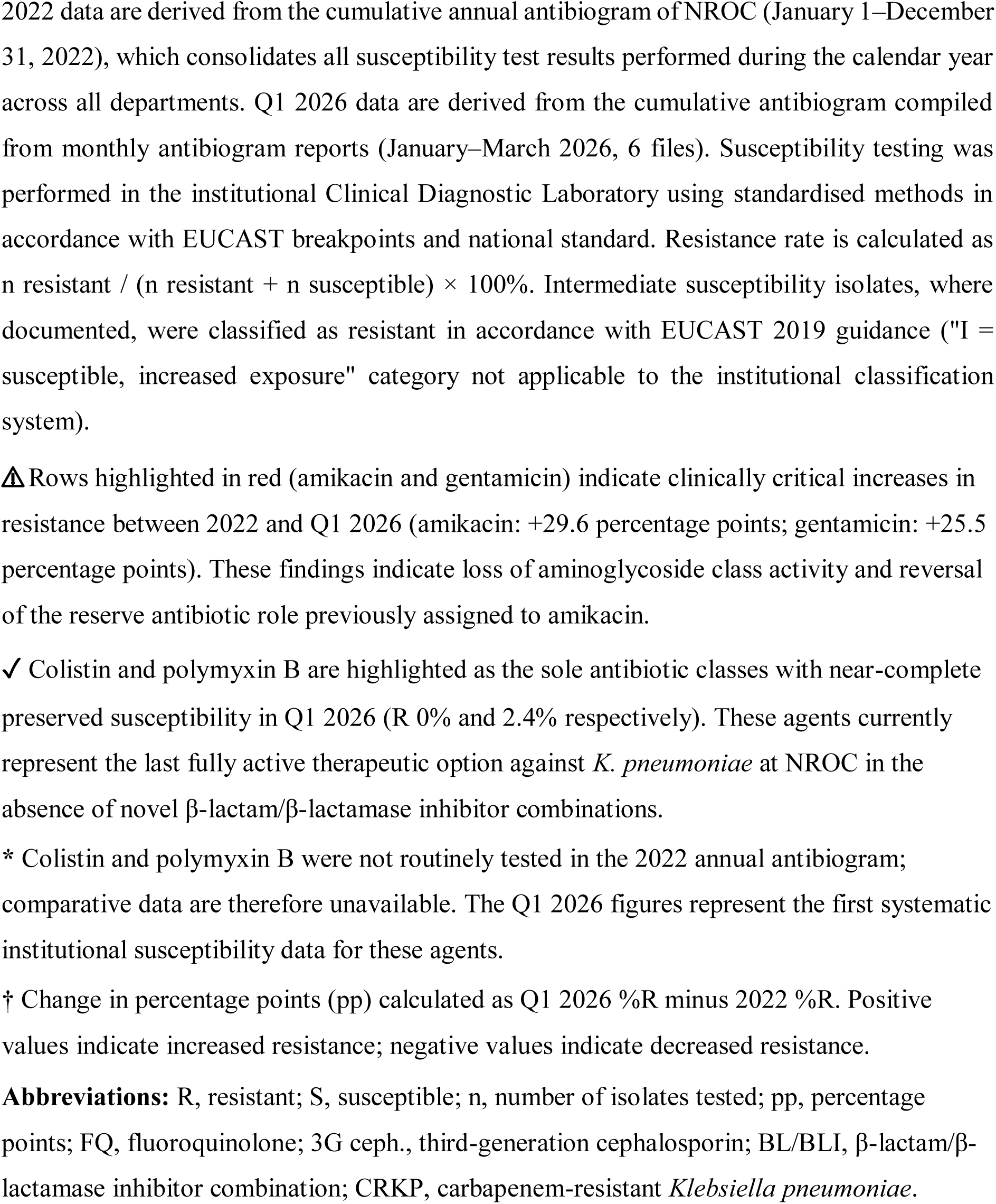
Antibiotic susceptibility of Klebsiella pneumoniae, NROC Astana: 2022 versus Q1 2026 (% resistant isolates)

| Antibiotic (class) | 2022 n tested | 2022 %R | 2026 n tested | 2026 %R | Change (pp) | Clinical significance |
| --- | --- | --- | --- | --- | --- | --- |
| Meropenem (carbapenem) | 159 | 72.3% | 50 | 64.0% | −8.3 | Partial improvement; still critically high |
| Imipenem (carbapenem) | 148 | 72.3% | 50 | 62.0% | −10.3 | Partial improvement; still critically high |
| Ertapenem (carbapenem) | 144 | 78.5% | 41 | 56.1% | −22.4 | Largest improvement; remains above 50% |
| Amikacin (aminoglycoside) | 141 | 8.5% | 42 | 38.1% | ↑ +29.6 | ⚠ Critical — former reserve agent, now unreliable |
| Gentamicin (aminoglycoside) | 147 | 48.3% | 42 | 73.8% | ↑ +25.5 | ⚠ Class-wide aminoglycoside resistance trend |
| Colistin (polymyxin) | — | — | 16 | 0% | — | ✓ Sole fully active class in 2026 |
| Polymyxin B | — | — | 41 | 2.4% | — | ✓ Sole fully active class in 2026 |
| Tigecycline | — | — | 23 | 52.2% | — | Moderate activity; potential adjunct in combinations |
| Ciprofloxacin (FQ) | 159 | 88.1% | 49 | 77.6% | −10.5 | Improvement; remains very high |
| Ceftriaxone (3G ceph.) | 130 | 99.2% | 42 | 92.9% | −6.3 | Essentially inactive |
| Ceftazidime (3G ceph.) | 137 | 93.4% | 49 | 93.9% | +0.5 | Essentially inactive |

2022 data are derived from the cumulative annual antibiogram of NROC (January 1–December 31, 2022), which consolidates all susceptibility test results performed during the calendar year across all departments. Q1 2026 data are derived from the cumulative antibiogram compiled from monthly antibiogram reports (January–March 2026, 6 files). Susceptibility testing was performed in the institutional Clinical Diagnostic Laboratory using standardised methods in accordance with EUCAST breakpoints and national standard. Resistance rate is calculated as $n \text{ resistant} / (n \text{ resistant} + n \text{ susceptible}) \times 100\%$ . Intermediate susceptibility isolates, where documented, were classified as resistant in accordance with EUCAST 2019 guidance ("I = susceptible, increased exposure" category not applicable to the institutional classification system).
△ Rows highlighted in red (amikacin and gentamicin) indicate clinically critical increases in resistance between 2022 and Q1 2026 (amikacin: +29.6 percentage points; gentamicin: +25.5 percentage points). These findings indicate loss of aminoglycoside class activity and reversal of the reserve antibiotic role previously assigned to amikacin.
✓ Colistin and polymyxin B are highlighted as the sole antibiotic classes with near-complete preserved susceptibility in Q1 2026 (R 0% and 2.4% respectively). These agents currently represent the last fully active therapeutic option against *K. pneumoniae* at NROC in the absence of novel β-lactam/β-lactamase inhibitor combinations.
\* Colistin and polymyxin B were not routinely tested in the 2022 annual antibiogram; comparative data are therefore unavailable. The Q1 2026 figures represent the first systematic institutional susceptibility data for these agents.
† Change in percentage points (pp) calculated as Q1 2026 %R minus 2022 %R. Positive values indicate increased resistance; negative values indicate decreased resistance.
**Abbreviations:** R, resistant; S, susceptible; n, number of isolates tested; pp, percentage points; FQ, fluoroquinolone; 3G ceph., third-generation cephalosporin; BL/BLI, β-lactam/β-lactamase inhibitor combination; CRKP, carbapenem-resistant *Klebsiella pneumoniae*.

#### Carbapenems

Meropenem resistance declined modestly from 72.3% (2022, n=159) to 64.0% (Q1 2026, n=50), imipenem from 72.3% to 62.0%, and ertapenem from 78.5% to 56.1% — a reduction of 22.4 percentage points for ertapenem, the largest improvement across the study period. These changes, while clinically relevant, do not restore carbapenems to a position of reliable empirical utility: resistance remains above 56% for all three agents.

#### Amikacin — critical loss of activity

Amikacin resistance increased from 8.5% in 2022 (n=141) to 38.1% in Q1 2026 institution-wide (n=42), and to 60% in the intensive care sector specifically (СИТ, n=10). This represents a rise of +29.6 percentage points over four years and constitutes the most clinically consequential finding of this study, as amikacin was the only antibiotic with preserved majority susceptibility in 2022. Gentamicin resistance showed a parallel increase from 48.3% to 73.8% (+25.5 pp), confirming a class-wide aminoglycoside resistance trend.

#### Polymyxins — sole class with fully preserved activity

Colistin resistance remained at 0% (n=16) and polymyxin B resistance at 2.4% (1/41 isolates) in Q1 2026. These agents now represent the only antibiotic class with near-complete preserved susceptibility against *K. pneumoniae* at NROC. Tigecycline resistance was 52.2% (12/23 isolates) — a moderate level that may support use in combination regimens for non-bacteraemic infections.

#### Other classes

Third-generation cephalosporins (ceftriaxone R 93%, ceftazidime R 94%), fluoroquinolones (ciprofloxacin R 78%), and penicillin combinations (amoxicillin/clavulanate R 98%, ampicillin R 100%) remain broadly inactive in 2026, consistent with 2022 patterns.

### 3.5 Longitudinal Antibiogram Trends (2019–2026)

The five-year institutional monitoring report (2019–2023) combined with the 2022 and Q1 2026 antibiograms enables an eight-year longitudinal view of key susceptibility trends (Table 5; Figure 1). Carbapenem susceptibility declined from 57% (meropenem/imipenem, 2019) to a nadir of 27% (2022) before showing partial recovery to 36–44% by Q1 2026. Ertapenem susceptibility declined continuously from 59% (2019) to 21% (2022–2023), with partial recovery to 44% in Q1 2026. Amikacin susceptibility, which remained high at 91–96% through 2019–2023, fell to 62% by Q1 2026, with the intensive care sector showing further deterioration to 40% (see Section 3.4).

**Table 5.**
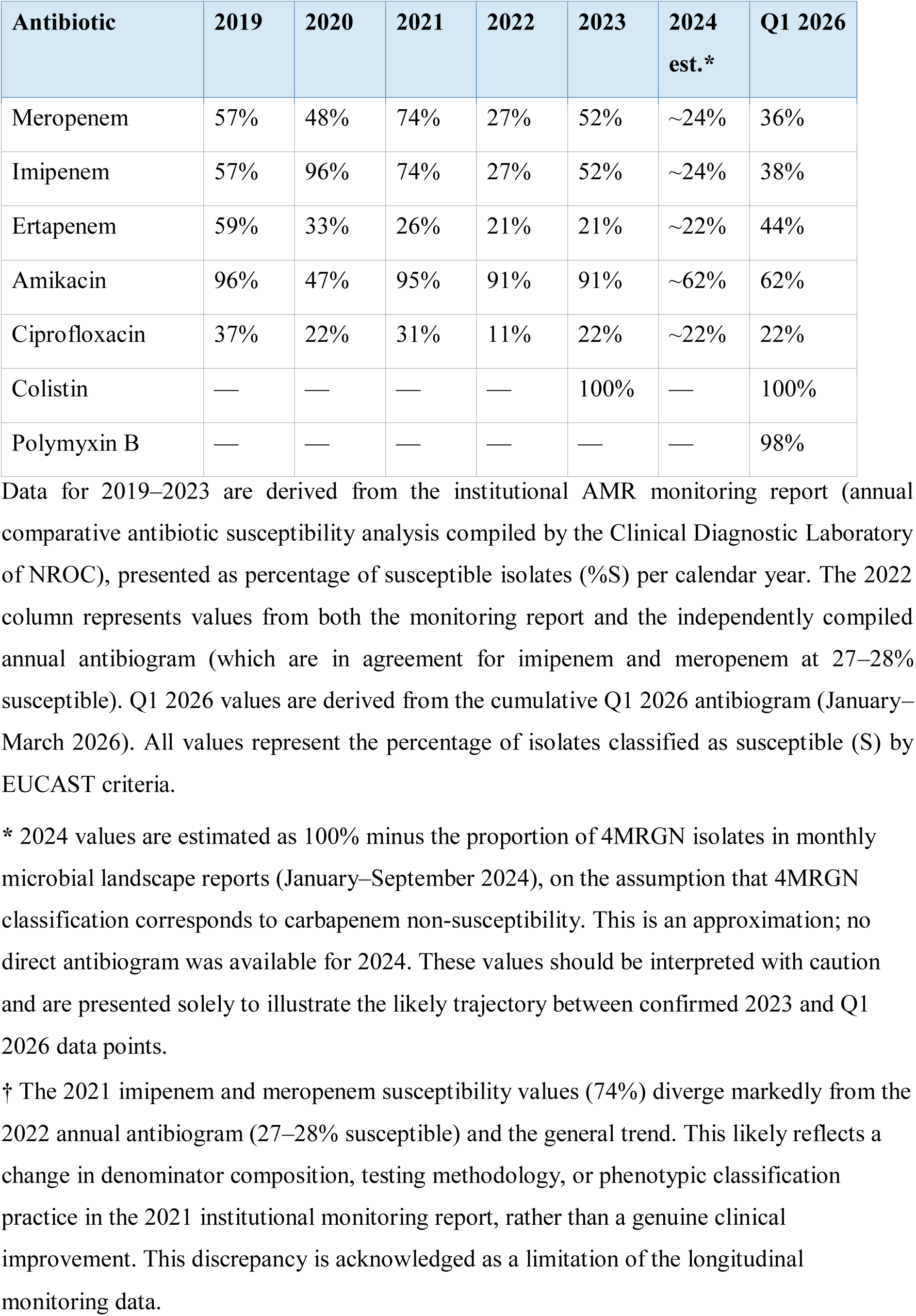
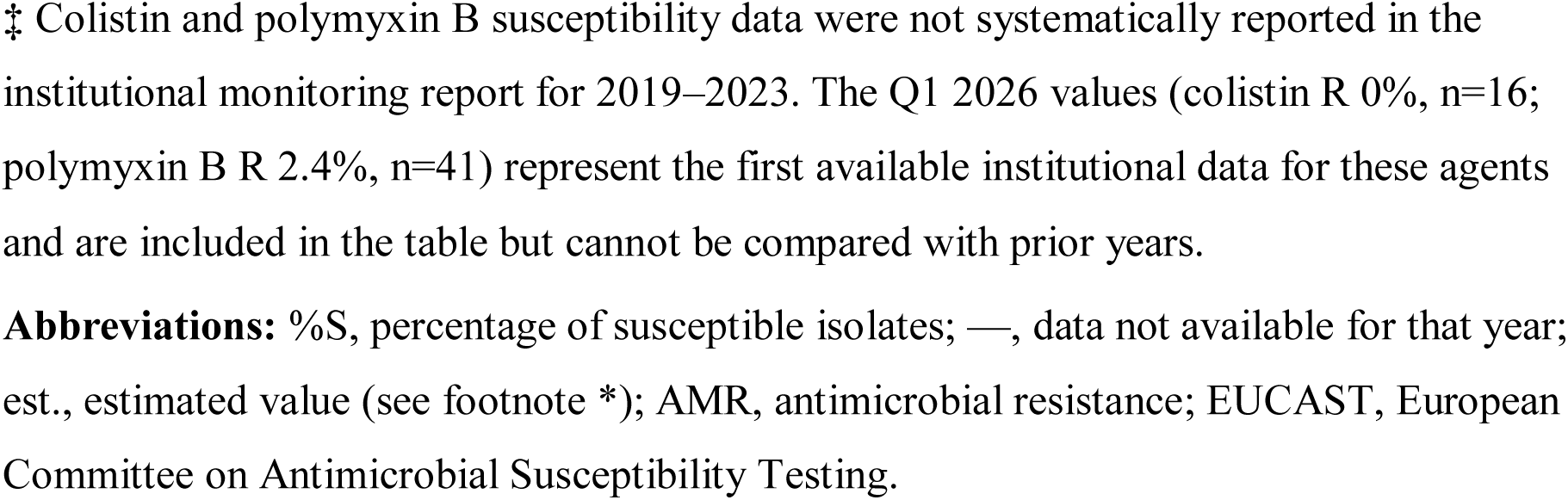
Longitudinal antibiotic susceptibility of Klebsiella pneumoniae at NROC, Astana, Kazakhstan, 2019–Q1 2026 (% susceptible isolates)

| Antibiotic | 2019 | 2020 | 2021 | 2022 | 2023 | 2024<br>est.* | Q1 2026 |
| --- | --- | --- | --- | --- | --- | --- | --- |
| Meropenem | 57% | 48% | 74% | 27% | 52% | ~24% | 36% |
| Imipenem | 57% | 96% | 74% | 27% | 52% | ~24% | 38% |
| Ertapenem | 59% | 33% | 26% | 21% | 21% | ~22% | 44% |
| Amikacin | 96% | 47% | 95% | 91% | 91% | ~62% | 62% |
| Ciprofloxacin | 37% | 22% | 31% | 11% | 22% | ~22% | 22% |
| Colistin | — | — | — | — | 100% | — | 100% |
| Polymyxin B | — | — | — | — | — | — | 98% |

Data for 2019–2023 are derived from the institutional AMR monitoring report (annual comparative antibiotic susceptibility analysis compiled by the Clinical Diagnostic Laboratory of NROC), presented as percentage of susceptible isolates (%S) per calendar year. The 2022 column represents values from both the monitoring report and the independently compiled annual antibiogram (which are in agreement for imipenem and meropenem at 27–28% susceptible). Q1 2026 values are derived from the cumulative Q1 2026 antibiogram (January–March 2026). All values represent the percentage of isolates classified as susceptible (S) by EUCAST criteria.
\* 2024 values are estimated as 100% minus the proportion of 4MRGN isolates in monthly microbial landscape reports (January–September 2024), on the assumption that 4MRGN classification corresponds to carbapenem non-susceptibility. This is an approximation; no direct antibiogram was available for 2024. These values should be interpreted with caution and are presented solely to illustrate the likely trajectory between confirmed 2023 and Q1 2026 data points.
† The 2021 imipenem and meropenem susceptibility values (74%) diverge markedly from the 2022 annual antibiogram (27–28% susceptible) and the general trend. This likely reflects a change in denominator composition, testing methodology, or phenotypic classification practice in the 2021 institutional monitoring report, rather than a genuine clinical improvement. This discrepancy is acknowledged as a limitation of the longitudinal monitoring data.
‡ Colistin and polymyxin B susceptibility data were not systematically reported in the institutional monitoring report for 2019–2023. The Q1 2026 values (colistin R 0%, n=16; polymyxin B R 2.4%, n=41) represent the first available institutional data for these agents and are included in the table but cannot be compared with prior years.
**Abbreviations:** %S, percentage of susceptible isolates; —, data not available for that year; est., estimated value (see footnote \*); AMR, antimicrobial resistance; EUCAST, European Committee on Antimicrobial Susceptibility Testing.

**Figure 1.**
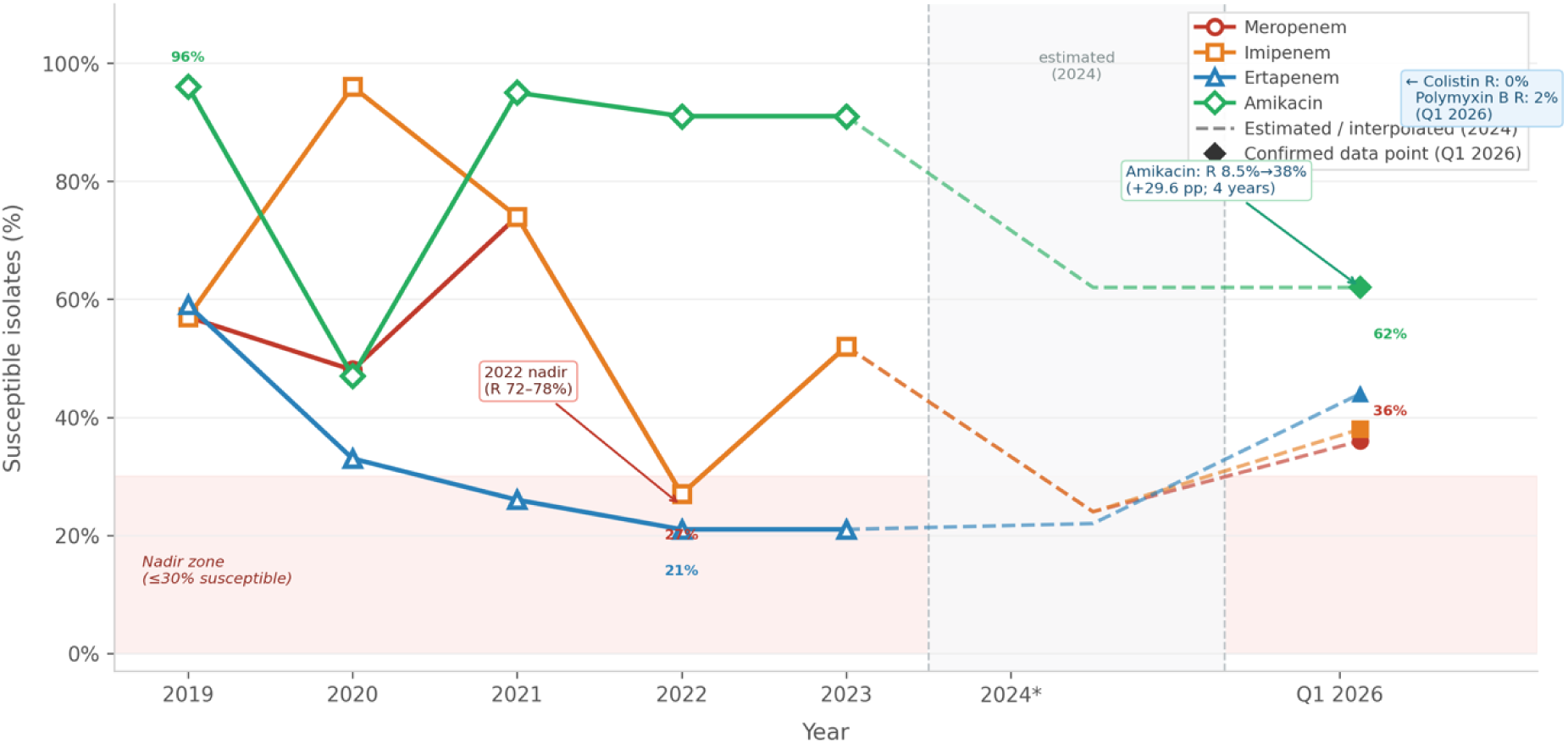
Longitudinal antibiotic susceptibility of Klebsiella pneumoniae at the National Research Oncology Centre (NROC), Astana, Kazakhstan, 2019–Q1 2026 (% susceptible isolates) The figure presents the percentage of susceptible (%S) isolates for four antibiotics representing critical therapeutic classes — meropenem, imipenem, ertapenem (carbapenems), and amikacin (aminoglycoside) — across an eight-year observation period. Solid lines with open markers (2019–2023) represent confirmed data points derived from the institutional AMR monitoring report (annual comparative antibiotic susceptibility analysis, Clinical Diagnostic Laboratory, NROC). Filled markers at Q1 2026 represent confirmed data from the institutional cumulative antibiogram for January–March 2026 (meropenem n=50; imipenem n=50; ertapenem n=41; amikacin n=42). The 2022 annual antibiogram independently corroborated the monitoring report values (meropenem %S 27.7%, n=159; amikacin %S 91.5%, n=141). Dashed lines connecting the 2023 and Q1 2026 data points pass through an estimated 2024 intermediate value, calculated as 100% minus the proportion of 4MRGN isolates in monthly microbial landscape reports (January–September 2024); these interpolated values are approximations and should not be interpreted as direct susceptibility measurements. The pink shaded band indicates the nadir susceptibility zone (≤30% susceptible isolates), corresponding to the period of maximum carbapenem resistance documented in the 2022 annual antibiogram (meropenem R 72.3%; imipenem R 72.3%; ertapenem R 78.5%). The grey shaded region with dashed vertical boundaries demarcates the estimated data period (2024). Three key trends are illustrated: (1) progressive decline and partial recovery of carbapenem susceptibility (meropenem/imipenem: 57%→27%→36–38%; ertapenem: 59%→21%→44%); (2) sustained erosion of amikacin susceptibility from 2023 to Q1 2026 (91%→62%), representing a net loss of 29–34 percentage points from the 2019 baseline of 96%; and (3) the contrast between the aminoglycoside decline and the relative stability of carbapenem susceptibility in the most recent period, suggesting differential selection pressure dynamics. Colistin (R 0%, n=16) and polymyxin B (R 2.4%, n=41) in Q1 2026 are not shown as line plots owing to the absence of prior-year comparator data; these agents currently represent the sole antibiotic class with near-complete preserved susceptibility against *K. pneumoniae* at NROC. **\*** 2024 tick label on the x-axis represents the estimated midpoint value (mid-2024), not a confirmed antibiogram data point. **†** The 2021 imipenem/meropenem susceptibility value (74%) likely reflects a change in denominator composition or classification practice rather than a genuine clinical improvement, and should be interpreted with caution (see Table 5, footnote †). **Abbreviations:** %S, percentage of susceptible isolates; R, resistant; n, number of isolates tested; AMR, antimicrobial resistance; CRKP, carbapenem-resistant *Klebsiella pneumoniae*; NROC, National Research Oncology Centre, Astana, Kazakhstan.

## 4. Discussion

### 4.1 CRKP as the Dominant Institutional Phenotype

The central and consistent finding across all periods of this study is that CRKP constitutes the dominant *K. pneumoniae* phenotype at NROC, with proportions of 62–76% across 2022–2026 and MDR rates of 76–87% throughout. This prevalence substantially and consistently exceeds the global pooled mean of 28.7%^[14]^ and is comparable only to the most severely affected settings globally: Greece (70.6%), India (67.6%), and Taiwan (67.5%).^[14]^ To our knowledge, this is the first systematic published documentation of this phenomenon from Kazakhstan or any Central Asian institution. The clinical implications are severe: 48% of all *K. pneumoniae* isolates in 2024 were from blood cultures, and 86.8% of these BSI isolates were CRKP — indicating a population facing a critical risk of lethal bloodstream infection with an organism against which standard antibiotics are broadly ineffective. The emergence of carbapenem-resistant hypervirulent strains (CR-hvKp) represents an additional and particularly lethal dimension of this threat, combining the resistance of CRKP with the virulence of hypervirulent lineages. ^[15, 16]^

### 4.2 The Progressive Loss of Amikacin: A Critical and Irreversible Turning Point

The most clinically consequential finding of this study is the progressive erosion of amikacin susceptibility between 2022 and 2026. In 2022, amikacin was the sole antibiotic class with preserved majority activity against *K. pneumoniae* at NROC, with a resistance rate of only 8.5% (12/141 isolates) — a finding that underpinned our previous recommendation of amikacin as the principal reserve agent in the local context of limited access to newer β-lactam/β-lactamase inhibitor (BL/BLI) combinations. By Q1 2026, institution-wide amikacin resistance had risen to 38.1% (16/42 isolates), and to 60% in the intensive care sector specifically — a net increase of 29.6–51.5 percentage points in four years.

This trajectory has several important implications. First, amikacin can no longer be considered a reliable empirical agent against *K. pneumoniae* at NROC: the probability of treatment failure with empirical amikacin monotherapy now exceeds 38% institution-wide and 60% in critical care. Second, the parallel increase in gentamicin resistance (48%→74%) confirms that this is a class-wide aminoglycoside resistance trend, not an amikacin-specific phenomenon — likely reflecting the widespread use of amikacin as a reserve agent over the preceding years, selecting for aminoglycoside-modifying enzyme production and 16S rRNA methylase acquisition. Third, this renders the antibiotic landscape at NROC significantly more constrained than four years ago: where amikacin provided a clinically accessible rescue option in 2022, no comparable option now exists outside polymyxins.

The loss of amikacin susceptibility is a direct and predictable consequence of deploying a reserve antibiotic without a stewardship programme to protect it. This finding should serve as a cautionary example and underscores why the protective stewardship of any remaining active agent — including polymyxins — is not optional but existentially urgent.

### 4.3 Polymyxins as the Last Active Class: Opportunities and Limits

The Q1 2026 antibiogram identifies colistin (R 0%, n=16) and polymyxin B (R 2.4%, n=41) as the only antibiotic classes with near-complete preserved activity against *K. pneumoniae* at NROC. This finding has both therapeutic implications and warnings. On the positive side, the preservation of polymyxin susceptibility provides a treatment anchor for life-threatening CRKP infections in the current absence of approved newer agents. Infectious Diseases Society of America (IDSA) 2024 guidance acknowledges the role of polymyxin-based combinations as salvage regimens when novel BL/BLI combinations are unavailable.^[17]^

However, polymyxins carry significant limitations that restrict their clinical utility. Nephrotoxicity is a major concern, occurring in 30–60% of patients, and is dose-limiting in patients with pre-existing renal impairment — a frequent characteristic in oncological and post-transplant populations.^[17]^ Polymyxin monotherapy is associated with poor microbiological outcomes and rapid resistance selection; combination with other agents (e.g., rifampicin, fosfomycin, or a carbapenem despite resistance) is generally recommended but has inconsistent evidence. Tigecycline, with 52% resistance in Q1 2026, may serve as a combination partner for non-BSI infections (its pharmacokinetics are unfavourable for bacteraemia), but the resistance rate is already concerning and warrants close monitoring.

The preservation of polymyxin susceptibility must now be treated as a critical institutional resource. Any uncontrolled or empirical use of colistin or polymyxin B risks accelerating resistance selection — potentially closing the last therapeutic window available for CRKP at NROC. Polymyxin use should be subject to mandatory pre-authorisation, microbiological confirmation, and de-escalation review within 48–72 hours.

### 4.4 Partial Carbapenem Recovery: Genuine Improvement or Survivor Bias?

The modest reduction in carbapenem resistance between 2022 and Q1 2026 (meropenem R 72%→64%; ertapenem R 78%→56%) is a genuinely interesting observation that warrants cautious interpretation. One potential explanation is that the amikacin use which was concentrated in ICU settings in 2022–2023 reduced exposure-based carbapenem selection, allowing a partial rebalancing of the phenotype distribution toward more susceptible variants. An alternative explanation is denominator change: the Q1 2026 dataset includes all departments including lower-acuity wards where susceptible strains predominate, which could dilute the apparent resistance rate relative to pure ICU data from 2022. A third possibility is a genuine clinical benefit from emerging infection control practices, which the institutional strategic plan documents as under development.

Regardless of mechanism, a meropenem resistance rate of 64% and ertapenem resistance of 56% do not restore carbapenems to clinical utility as empirical agents. For targeted therapy of confirmed susceptible isolates they remain an option, but the probability of susceptibility in any unselected *K. pneumoniae* isolate at NROC is below 40% — insufficient for reliable empirical use in critically ill patients.

### 4.5 The 2025–2026 Pathogen Shift: Broader Enterobacterales Crisis

Global surveillance data confirm that both AMR and virulence determinants in K. pneumoniae are increasing in prevalence across host sources worldwide.^[18]^ Beyond *K. pneumoniae* itself, the concurrent emergence of MDR *Escherichia coli* (ESBL+, MDR) as the dominant pathogen in ТКМ and surgical ICU departments in 2025–2026 signals that the AMR crisis at NROC has expanded to a broader Enterobacterales resistance problem. This shift is a well-recognised consequence of sustained selective pressure from carbapenems and CRKP-directed therapy: heavy carbapenem use selects for ESBL-producing *E. coli* and other Enterobacterales that were suppressed when carbapenems were broadly effective.^[19]^ The emergence of MDR *E. coli* as dominant in HSCT settings is particularly alarming given the severity of BSI in neutropenic patients and the increasingly limited coverage options.

### 4.6 Haemato-oncological and Transplant Settings as Epicentres

The bone marrow transplantation centre displayed consistently extreme MDR rates across all periods (86–98%), along with a >2.5-fold increase in total clinical isolates between 2024 (n≈45) and 2025 (n=118). This exponential growth in microbiological burden in the highest-risk patient population is the most alarming institutional trend documented in this study. HSCT recipients face profound and prolonged immunosuppression, repeated invasive procedures, and extended exposure to broad-spectrum antibiotics — all established drivers of MDR acquisition and infection.^[14]^ Dedicated CRKP and MDR Enterobacterales colonisation screening at HSCT admission, active contact precautions for carriers, and consideration of selective digestive decontamination protocols are urgently indicated.

### 4.7 Limitations

This study has several limitations. First, all data originate from a single tertiary oncology centre; results are not generalisable to other Kazakhstani or Central Asian institutions. Second, data coverage is incomplete for some periods: full-year 2022 landscape data are unavailable (only Q4 captured), and 2023 landscape data cover only January–August. Third, the 2025 data source (clinical register) excludes routine surveillance cultures, rendering it non-comparable in denominator to landscape reports. Fourth, the Q1 2026 antibiogram covers only 50 isolates over three months, limiting statistical power for between-period comparisons. Fifth, the dataset provides aggregate isolate counts without patient-level linkage, precluding outcome analysis, risk factor identification, duplicate exclusion, or treatment response assessment. Sixth, no carbapenemase genotyping (KPC, NDM, OXA-48 etc.) was performed, limiting molecular epidemiological conclusions. Whole-genome sequencing-based (WGS) surveillance would enable identification of dominant sequence types and transmission clusters, as the global population structure of K. pneumoniae is dominated by a small number of high-risk clonal lineages.^[20]^ WGS studies are needed to identify dominant clones, characterise resistance mechanisms, and investigate transmission networks.

## 5. Conclusions

This four-year surveillance study provides the first systematic AMR evidence from a Kazakhstani tertiary centre. CRKP dominated the *K. pneumoniae* phenotype throughout 2022–2026 (62–76%), with MDR rates 2–3 times above global averages — an established institutional reality, not an emerging threat. Amikacin resistance rose from 8.5% to 38–60% over four years, leaving polymyxins as the sole active class; this loss demonstrates the cost of deploying reserve antibiotics without stewardship. Concurrently, MDR *E. coli* displaced CRKP as the dominant pathogen in key departments, and aggregate MDR burden continued to rise. Five urgent actions are required: (1) formal antimicrobial stewardship programme with carbapenem and polymyxin pre-authorisation; (2) active CRKP colonisation screening and contact precautions in ТКМ and ICU settings; (3) formulary introduction of novel BL/BLI combinations (ceftazidime-avibactam, meropenem-vaborbactam, or cefiderocol); (4) carbapenemase genotyping for targeted therapy and outbreak detection; and (5) integration into national AMR reporting aligned with WHO GLASS. Central Asia remains a critical surveillance blind spot — this study demonstrates both the severity of the problem and the feasibility of addressing it using existing laboratory infrastructure.

## Data Availability

All data produced in the present study are available upon reasonable request to the authors

## Declarations

### Ethics approval and consent to participate

This study was approved by the Local Ethics Committee of the National Centre for Biotechnology LLP, Astana, Kazakhstan (approval date: October 8, 2025). The study was conducted in accordance with the Declaration of Helsinki. All microbiological data were collected as part of routine institutional clinical practice and anonymised prior to analysis. The requirement for individual patient informed consent was waived by the Ethics Committee in accordance with national regulatory provisions for retrospective and prospective analysis of anonymised administrative and laboratory records

### Consent for publication

Not applicable.

### Availability of data and materials

The datasets used and/or analyzed during the current study are available from the corresponding author upon reasonable request

### Authors’ contributions

GAY: study design, data collection, data analysis and interpretation, manuscript writing and revision. IYM: manuscript revision and critical review. YVZ: data analysis and interpretation, manuscript revision. YAM: data collection. YSA: data collection. GSB: data collection. SBS: data collection. MKH: data collection. ZAB: study supervision, manuscript revision. All authors revised the drafted manuscript, and all read and approved its final version.

## Acknowledgements

The authors wish to thank the staff of the Clinical Diagnostic Laboratory of the National Research Oncology Centre, Astana, Kazakhstan, for their sustained commitment to institutional microbiological surveillance and for maintaining the laboratory records that made this study possible. The authors also thank the infection control team at NROC for their collegial support during data collection.

## List of Abbreviations

AMR: antimicrobial resistance
APC: article processing charge
BAL: bronchoalveolar lavage
BSI: bloodstream infection
BL/BLI: β-lactam/β-lactamase inhibitor combination
CRKP: carbapenem-resistant *Klebsiella pneumoniae*
ESBL: extended-spectrum β-lactamase
EUCAST: European Committee on Antimicrobial Susceptibility Testing
FQ: fluoroquinolone
GBD: Global Burden of Disease
GLASS: Global Antimicrobial Resistance and Use Surveillance System (WHO)
HSCT: haematopoietic stem cell transplantation
ICU: intensive care unit
IDSA: Infectious Diseases Society of America
KPC: Klebsiella pneumoniae carbapenemase
MDR: multidrug-resistant
NDM: New Delhi metallo-β-lactamase
NROC: National Research Oncology Centre, Astana, Kazakhstan
OXA-48: oxacillinase-48 carbapenemase
3MRGN: 3-class multidrug-resistant Gram-negative organism (carbapenem-susceptible)
4MRGN: 4-class multidrug-resistant Gram-negative organism (carbapenem-resistant; equivalent to CRKP in this study)
UTI: urinary tract infection
WGS: whole-genome sequencing
WHO: World Health Organization

